# High-throughput splicing assays identify missense and silent splice-disruptive *POU1F1* variants underlying pituitary hormone deficiency

**DOI:** 10.1101/2021.02.04.21249469

**Authors:** Peter Gergics, Cathy Smith, Hironori Bando, Alexander A. L. Jorge, Denise Rockstroh-Lippold, Sebastian Vishnopolska, Frederic Castinetti, Mariam Maksutova, Luciani Renata Silveira Carvalho, Julia Hoppmann, Julian Martinez Mayer, Frédérique Albarel, Debora Braslavsky, Ana Keselman, Ignacio Bergadá, Marcelo Martí, Alexandru Saveanu, Anne Barlier, Rami Abou Jamra, Michael H. Guo, Andrew Dauber, Marilena Nakaguma, Berenice B Mendonça, A Bilge Ozel, Qing Fang, Qianyi Ma, Jun Z. Li, Thierry Brue, María Ines Pérez Millán, Jacob O. Kitzman, Ivo JP Arnhold, Roland Pfaeffle, Sally A. Camper

## Abstract

Pituitary hormone deficiency occurs in ∼1:4,000 live births. Approximately 3% of the cases are due to mutations in the alpha isoform of POU1F1, a pituitary-specific transcriptional activator. We found four separate heterozygous missense variants in unrelated hypopituitarism patients that were predicted to affect a minor isoform, POU1F1 beta, which can act as a transcriptional repressor. These variants retain repressor activity, but they shift splicing to favor the expression of the beta isoform, resulting in dominant negative loss of function. Using a high throughput splicing reporter assay, we tested 1,080 single nucleotide variants in *POU1F1*. We identified 113 splice disruptive variants, including 23 synonymous variants. We evaluated separate cohorts of hypopituitarism patients and found two different synonymous splice disruptive variants that co-segregate with hypopituitarism. This study underlines the importance of evaluating the impact of variants on splicing and provides a catalog for interpretation of variants of unknown significance in the *POU1F1* gene.

## Introduction

*POU1F1* (formerly PIT-1) is a signature pituitary transcription factor that directly regulates the transcription of growth hormone (*GH)*, prolactin (*PRL)*, and both the alpha (*CGA*) and beta (*TSHB*) subunits of thyroid stimulating hormone^1,2^. In mice, *Pou1f1* is expressed after the peak expression of *Prop1* at E14.5 and remains expressed into adulthood^3,4^. A well-characterized mutant of *Pou1f1* (*Pit1*^*dw/dw*^) carries a spontaneous missense mutation (p.W251C) in the homeodomain that disrupts DNA binding^4,5^. The homozygous mutant mice have no somatotrophs, lactotrophs or thyrotrophs except for the *Pou1f1*-independent rostral tip thyrotrophs^4,6-8^. In humans, loss of *POU1F1* function typically results in GH, TSH and PRL deficiency^9^.

*POU1F1* undergoes an evolutionarily conserved program of alternative splicing^10,11^, resulting in a predominant isoform, alpha, that acts as a transcriptional activator and a minor isoform, beta, that acts as a transcriptional repressor^12-14^. In the human pituitary gland, the beta isoform comprises approximately 1-3% of *POU1F1* transcripts ^10,15^. The POU1F1 beta isoform transcript is created by utilization of an alternative splice acceptor sequence for exon 2, located 78 bp upstream of the alpha acceptor, resulting in a 26 amino acid insertion that encodes an ETS1 binding domain. This insertion, which is absent in the alpha isoform, disrupts the transactivation domain at amino acid 48. The POU1F1 alpha and beta isoforms have different activities depending on the context of the target gene^12^. For example, the POU1F1 alpha isoform activates its own expression, but the beta isoform does not, and the beta isoform interferes with alpha isoform mediated activation^14^.

The first case of a recessive *POU1F1* loss of function was described in a patient with CPHD born to consanguineous parents^16^, and since then thirty-seven unique variants in *POU1F1* have been reported in patients with CPHD or IGHD^17-23^. A few dominant negative mutations have been reported: p.P76L alters the transactivation domain and causes completely penetrant IGHD^24^, p.K216E interferes with the ability of POU1F1 to interact with retinoic acid receptors and p300^25^, and p.R271W interferes with the ability of POU1F1 to be tethered to the nuclear matrix through MATR3, SATB1 and CTNNB1^26^. All of the reported mutations are located in domains shared by the alpha and beta isoforms of POU1F1 and were functionally tested using the alpha isoform only.

We found four missense variants, in four independent families, that shift splicing to favor the POU1F1 beta isoform almost exclusively, while retaining its transcriptional repressor activity on the *POU1F1* enhancer. We used a high throughput assay to identify 113 variants in and around exon 2 that cause exon skipping, cryptic splicing and isoform switching. We used this catalog to evaluate additional families with hypopituitarism and identified two unrelated patients carrying synonymous *POU1F1* variants that affect its splicing without changing the amino acid sequence. This study underscores the importance of evaluating splicing defects as a disease mechanism.

## Methods

### Patients

The studies were approved by ethical committees: the local Comite de Eticae Pesquisa da Faculdade de Medicina da Universidade de São Paulo (CEP-FMUSP) and the national Comite nacional de etica em pesquisa (CONEP) CAAE, 06425812.4.0000.0068; the Ethics Committee of the Faculty of Medicine, University of Leipzig (UL), Karl-Sudhoff-Institute for Medical History and Natural Sciences, Käthe-Kollwitz-Straße 82, 04109 Leipzig, Germany; and the Comité de Ética en Investigación (Research Ethics Committee) of the Hospital de Niños Ricardo Gutierrez (HNG), Gallo 1330, Ciudad autónoma de Buenos Aires, Argentina (CEI N° 16.06). The GENHYPOPIT network collected anonymized information in a database declared to health authorities in accordance with local regulations at Aix-Marseille Université (AMU) - Conception Hospital (Assistance Publique - Hôpitaux de Marseille, AP-HM), and a declaration was made to the National Commission for Data Protection and Liberties (CNIL-France): 1991429 v 0. Patients or their parents signed a written informed consent to participate. Families 1, 3, and 6 are historical cases that were referred to the GENHYPOPIT network for genetic testing. Limited information is available for Families 1 and 3, and they were lost for follow up. The University of Michigan Institutional Review Board (UM) found the study exempt because patient DNA samples were anonymized before exome sequencing at UM.

### DNA sequencing of patient samples

Individuals from Families 1, 2, 4, and 5 underwent whole exome sequencing (WES). Representative *POU1F1* variants in Family 3 and 6 were discovered in a traditional CPHD candidate gene screening using Sanger sequencing (*PROP1, POU1F1, LHX3* and *LHX4*). WES of Families 1 and 5 was carried out at University of Michigan as previously described^17^. WES of Family 2 was performed at the Broad Institute as previously described^27^. WES of Family 4 was performed at the Institute of Human Genetics at University of Leipzig.

### Expression vectors and cell culture

The open reading frame of either *POU1F1* isoform alpha (NM_000306.3) or beta (NM_001122757.2) was cloned into pcDNA3.1+/C-(K)-DYK. Site directed mutagenesis was used to obtain each of the variant POU1F1 beta isoforms: p.S50A, p.I51S, p.L52W, and p.S53A (Genscript). A firefly luciferase reporter gene was constructed in pNBm81-luc with 14 kb of the mouse *Pou1f1* 5’ flanking sequences that includes early and late enhancers and the promoter, and 13 bp of the 5’UTR. Cloning was performed with Infusion HD (Clontech) or NEBuilder HiFi DNA Assembly (New England Biolabs). Plasmid sequences were confirmed by Sanger sequencing. The pRL-TK renilla (Promega) was used as a normalization control and pcDNA3.1(-) (Thermo-Fisher) to keep the total DNA constant. COS-7 cells were purchased from the American Type Culture Collection. Cells were maintained in Dulbecco’s modified eagle medium (DMEM, Gibco, Grand Island, NY, USA) containing 10% fetal bovine serum and pen-strep (Gibco). Plasmids were transiently transfected into COS-7 cells using ViaFect Transfection Reagent (Promega, Madison, WI, USA). Luciferase activities were measured as suggested by the manufacturer (Dual-luciferase assay system; Promega).

### Exon trapping assay

Human *POU1F1* exon 2, flanked by partial intron 1 (85 bp upstream) and intron 2 (178 bp downstream), was cloned into the *BamHI* cloning site of the pSPL3 plasmid (Invitrogen) to create an exon trapping plasmid with a total insert size of 413 bp. Similarly, a minigene exon trapping plasmid was constructed that included the last 85 bp of intron 1 and the first 85 bp of intron 5, for a total insert size of 3,442 bp including exons 2, 3, 4, and 5. Site directed mutagenesis was used to create the desired variants. Plasmids were transiently transfected into COS-7 cells. Total RNA was purified with RNeasy mini (Qiagen). After reverse transcription, we analyzed exon trapping using RT-PCR with following primers; Primers SD6 Forward (5’-TCT GAG TCA CCT GGA CAA CC-3’) and SA2 reverse (5’-ATC TCA GTG GTA TTT GTG AGC -3’)^28^.

### *POU1F1* Saturation Mutagenesis

The cloned *POU1F1* fragment in pSPL3 was divided into four overlapping tiles of 150 bp each, spanning exon 2 plus flanking introns (79 bp upstream to 131 bp downstream). Mutant tile libraries containing every possible single nucleotide variant were synthesized as a single 150mer oligonucleotide pool by Twist Bio. HiFi Assembly was used to replace each wild type tile with the respective mutant tile library amplified from the oligo pool. The resulting mutant minigene library pools were transformed in 10b *E. coli* (New England Biolabs), with a minimum coverage of 90 clones per mutation.

### Mutant library barcoding and sequencing

To tag each mutant minigene clone with a unique barcode, a random barcode sequence (N_20_) was inserted by HiFi Assembly into the MscI site within the common 3’ UTR. Subassembly sequencing^29^ was used to pair each 3’ UTR barcode with its linked variant(s) in cis. Briefly, a fragment starting with the POU1F1 insert and ending at the N_20_ barcode (2.2 kb downstream) were amplified from the plasmid library DNA by PCR using 5’-phosphorylated primers. The resulting linear fragment was re-circularized by intramolecular ligation using T4 DNA ligase (NEB), to bring each barcode in close proximity to the mutagenized region. From this re-circularized product, paired-end amplicon sequencing libraries were generated, such that each reverse read contained a plasmid barcode and the paired forward read contained a sequence from the associated POU1F1 insert. Barcode reads were clustered with starcode^30^ (arguments “-d 1 -r 3”) to generate a catalog of known barcodes. Variants were called within each barcode group using freebayes^31^ and filtered to require majority support, and read depth _≥_4 along the entire region targeted for mutagenesis.

### Pooled exon-trap transfection and RNA-seq

COS-7 cells were plated at 5×10^6^ cells/60 mm plate. Each was transfected with 4 ug of the barcoded mutant exon-trap library using ViaFect reagent (3:1 ratio to DNA). After 24 hrs, RNA was purified as above, and 5 ug of total RNA was used to prepare first-strand cDNA using the SuperScript III First-Strand Synthesis kit (Invitrogen) with oligo dT primers. Spliced transcript was amplified using nested PCR, initially for 6 cycles using the SD6F/SA2R primers, followed by 20 cycles using primers SD2F/jklab0046 (TGTAGTCAGTGCCATCTTGGATCT). Paired-end Illumina sequencing libraries were generated by tailing PCR (6 cycles) with a forward primer within the constant upstream exon (GTGACTGGAGTTCAGACGTGTGCTCTTCCGATCT AGGGCATAGTGCCATCTTGGATCT) and a reverse primer immediately downstream of the N_20_ barcode (TCGTCGGCAGCGTCAGATGTGTATAAGAGACAGAGTGAACTGCACTGTG ACA AGCTGC). Unique dual i5/i7 indices were added by a second round of tailing PCR (6 cycles), and the resulting products were purified by SPRI bead cleanup and submitted for Illumina sequencing on a Hiseq 4000 and/or Novaseq instrument.

### RNA-seq processing pipeline

Reverse reads containing the plasmid barcode were searched for exact match to a known barcode from the plasmid library. Forward reads containing the spliced sequence were mapped to a variant-specific reference consisting of the POU1F1 exon trap reference sequence with the respective mutation introduced in silico, using GMAP^32^ (arguments “-t 8 -f samse --microexon-spliceprob=1.0 --allow-close-indels=2”). From the spliced reads, an isoform catalog was tallied requiring each isoform to be represented by at least three distinct barcodes and nine reads. Spliced reads associated with each barcode were tallied to produce per-barcode isoform usage counts, and percent spliced in (PSI) fractions. Barcodes corresponding to the same *POU1F1* variant were then aggregated (weighted by the number of reads obtained) to generate for each variant a mean PSI score for all known isoforms. Isoforms not matching a known isoform (beta, skip, or alpha) were placed in a catch-all category called “OTHER”. Barcodes represented by fewer than three reads were discarded from further analyses. Then, within each biological replicate, the mean PSI score was standardized, and variants with a *z*-score ≥ 1.5 for the beta, skip, or other isoform in at least half of the replicates were nominated as splice disruptive variants.

### Comparison of bioinformatic predictors

HAL delta_psi scores^33^, SPANR zdelta_psi scores^34^, SpliceAI ds_max scores^35^, and MMSplice delta_logit_psi scores^36^ were obtained from their original publications without modification. To compute per-variant ΔESRseq scores^37^, we took the difference between the mean ESRseq z-scores of hexamers overlapping a variant position from that of hexamers overlapping the corresponding wildtype position. Precision-recall curves were obtained to summarize each algorithm’s ability to predict the experimental determination of splice disruptiveness. For algorithms which output signed scores, area under the curve (prAUC) was separately computed using signed and absolute scores as input and the higher prAUC was taken.

### Selection of candidate RNA binding proteins (RBP)

RNACompete z-scores ^38^ were obtained from the cisBP-RNA database (http://cisbp-rna.ccbr.utoronto.ca). At each position, wild-type and variant-containing z scores were taken as the maximum among the overlapping kmers, and the difference taken between the wild-type and variant scores. Motifs with high scoring matches (wildtype z>=3) to the wild-type sequence in the beta variant cluster (c.143-1 to c.167) were then pursued further.

### Data availability

Raw sequence reads and processed counts are available at GEO (accession tbd). Jupyter notebooks to reproduce the processed dataset from raw counts are posted at GitHub (URL tbd).

## Results

### Mutations in the POU1F1 beta coding region cause hypopituitarism

We initially focused on four cases of hypopituitarism from different cohorts in Europe and South America (**Fig. 1A**). Affected individuals’ presentation was variable, ranging from multiple hormone deficiency with pituitary stalk interruption (Family 1) to isolated GH deficiency (Family 2) (**Table 1, Suppl. Fig. 1)**. The affected individuals had severe short stature and responded well to GH therapy (**Fig. 1B**). To identify causal variants, we performed whole exome sequencing (WES) for individuals in three families. Combined with conventional Sanger sequencing in another family, this revealed four missense variants in exon 2 of the *POU1F1* beta isoform, each in an unrelated family (**Figure 1A, 2A**). The four patient *POU1F1* missense variants are absent from gnomAD and in-house population-matched exome databases^39,40^, and they are predicted to be damaging by several bioinformatic algorithms (**Table 1**). Remarkably, these variants clustered in four consecutive codons: c.148T>G (p.S50A), c.152T>G (p.I51S), c.155T>G (p.L52W), and c.157T>G (p.S53A) in NM 001122757.3 (**Table 1**). Only one of these (c.155T>G, Family 3) appears to be *de novo*; the others were dominantly inherited and co-segregate with hypopituitarism phenotypes, except for c.148T>G which was inherited from the apparently unaffected parent in Family 1, indicating that if causal, this variant is incompletely penetrant. The other parent in Family 1, the two affected children, and one unaffected relative also carried a variant of uncertain significance, *SIX3* p.P74R. No other variants in known hypopituitarism genes were detected.

**Table 1.**
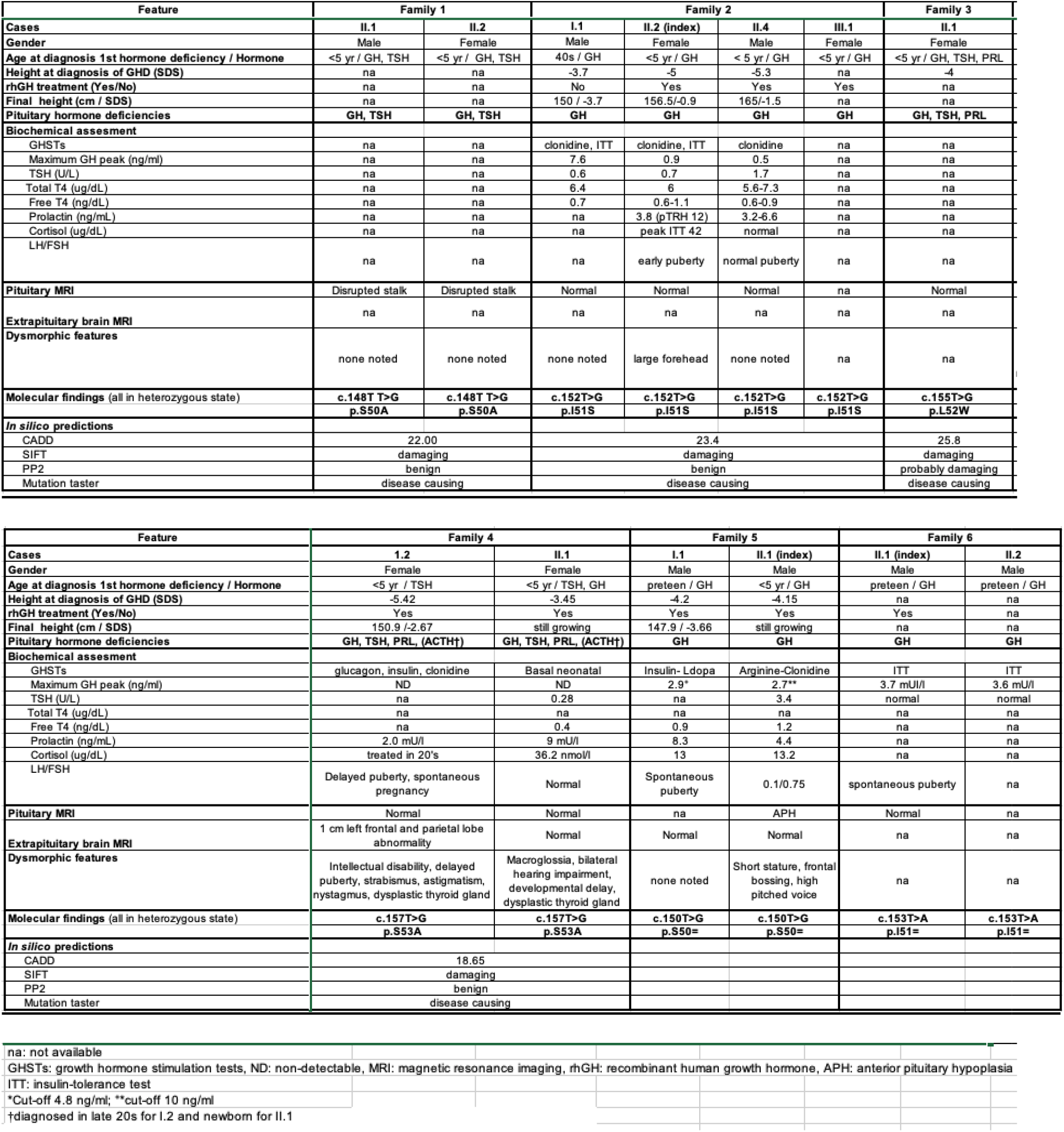
Clinical and molecular features of affected individuals

**Figure 1.**
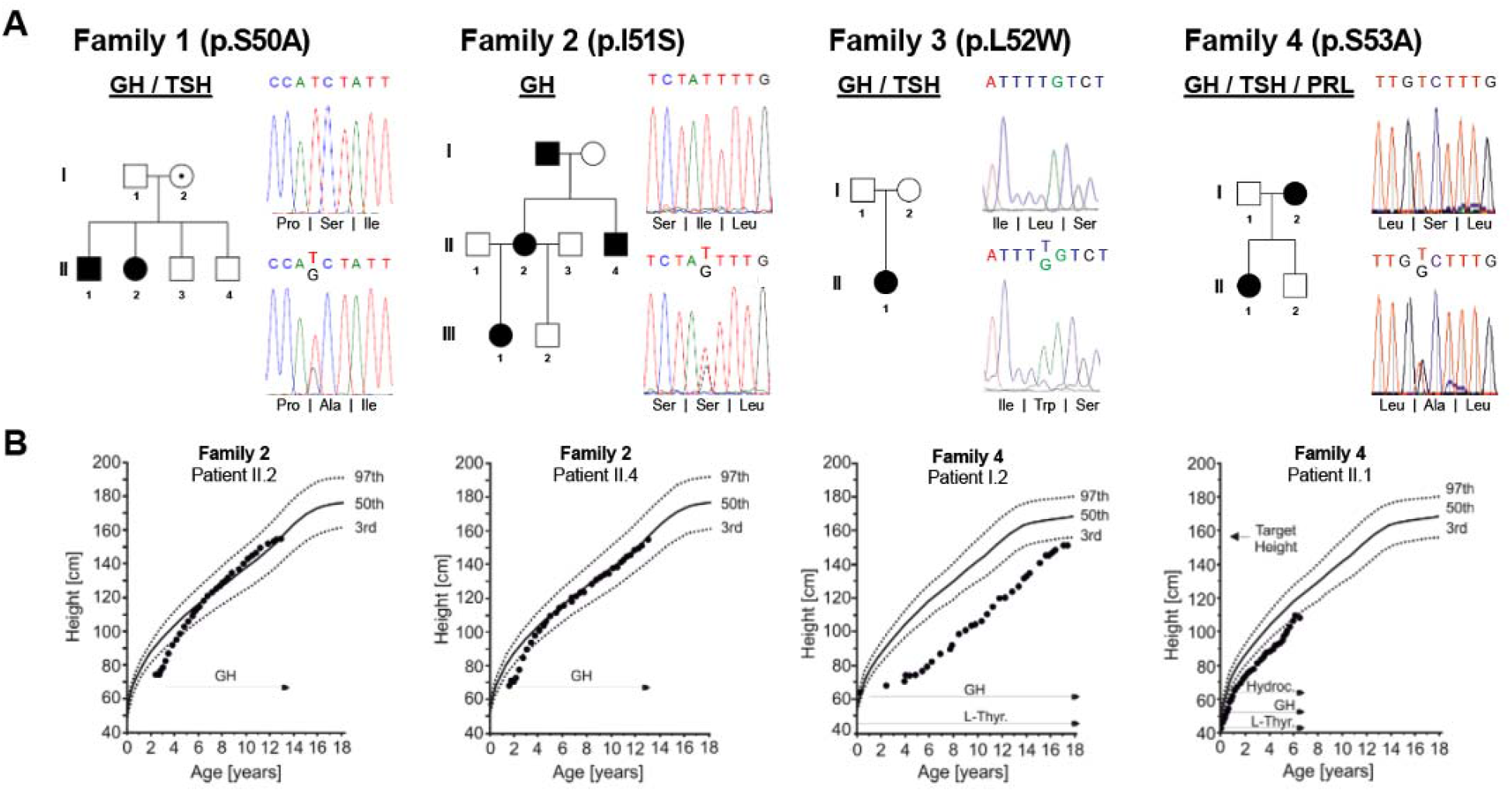
Clinical characteristics of the variants of POU1F1 beta coding region. **A**. Pedigrees and the sequence of the patients. Family 1-4 have variants in the POU1F1 beta coding region: c.148T>G (p.S50A), c.152T>G (p.I51S), c.155T>G (p.L52W), and c.157T>G (p.S53A). **B**. Growth curve of the patients from Family 2 and 4. GH replacement therapy was effective in reaching ideal height.

### Sequence variants retain POU1F1 beta isoform repressor function

We first asked whether these variants disrupt the ability of POU1F1 to bind and transactivate its own promoter^41^ using a transient transfection assay (**Figure 2B**). As expected, a *Pou1f1* promoter reporter was strongly activated when co-transfected with cDNA of POU1F1 alpha isoform, which does not include the variant sites. Neither WT POU1F1 beta isoform, nor any of the four patient missense variants, showed significant activation of the *Pou1f1*-luc reporter. Consistent with a repressive role for POU1F1 beta, co-transfection with alpha at a 1:1 ratio significantly suppressed activation compared to the equivalent amount of alpha isoform alone. The four POU1F1 beta variants and WT beta repressed POU1F1 alpha activity to a similar degree.

**Figure 2.**
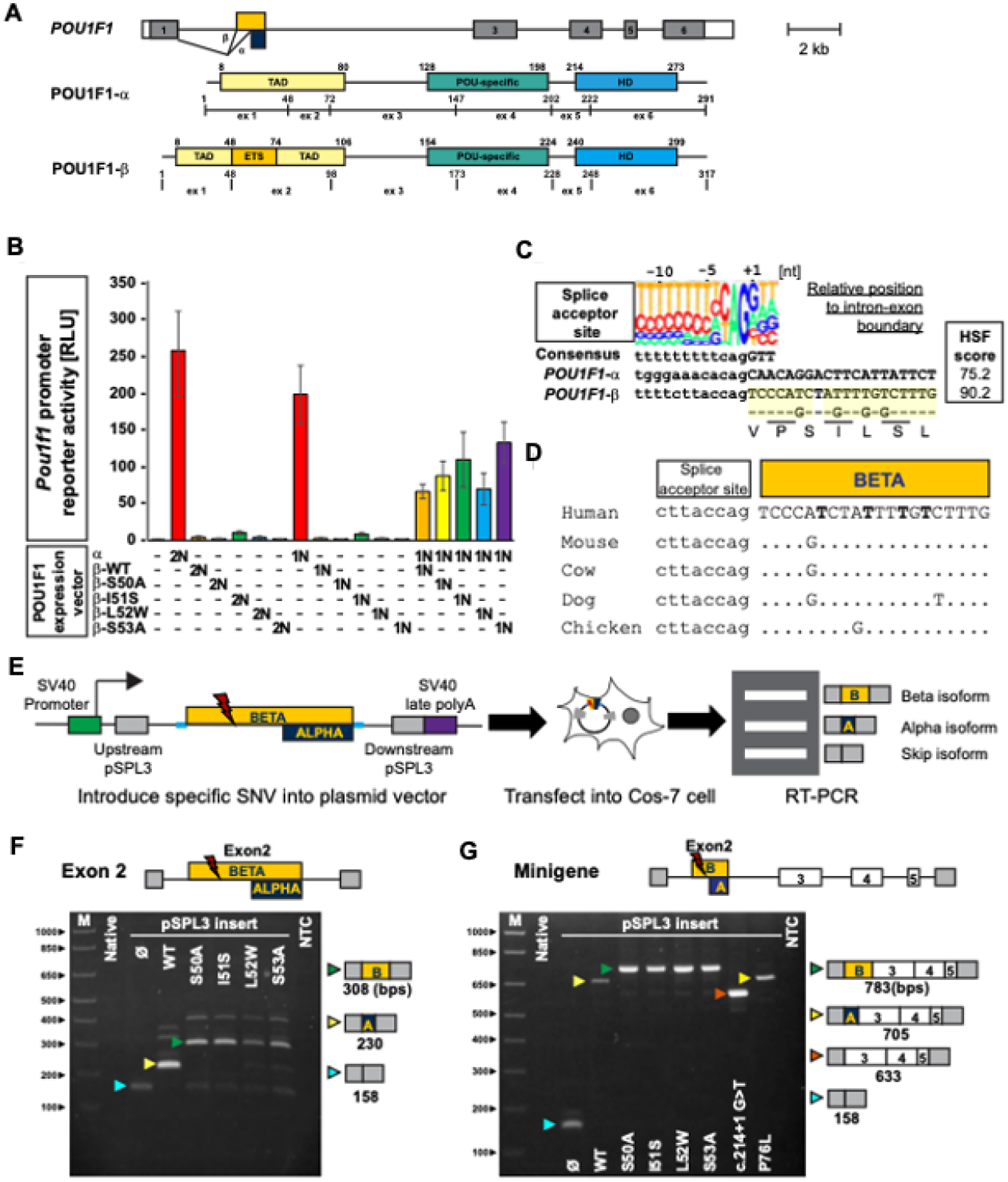
Variants in the *POU1F1* beta coding region suppress the function of alpha isoform and lead to splicing abnormality. **A**. Schematic of the human *POU1F1* gene and protein isoforms produced by use of alternate splice acceptors at exon 2. The *Pou1f1* beta isoform has an insertion of 26 amino acids located at amino acid 48 in the transactivation domain. **B**. COS-7 cells were transfected with a *Pou1f1-* luciferase reporter gene and expression vectors for POU1F1 alpha or beta isoforms either singly or together in the ratios indicated (2N and 1N). WT POU1F1 alpha has strong activation at 2N and 1N dosages. WT and variants of POU1F1 beta isoform have no significant activation over background. A 50:50 mix of alpha and WT beta isoforms exhibited reduced activation. The variant beta isoforms suppress alpha isoform mediated activation to a degree similar to WT. **C**. Diagram of the splice acceptor site consensus and the genomic DNA sequence at the boundary between intron 1 and splice sites utilized in exon 2 of the *POU1F1* gene^71^ **D**. Evolutionary conservation of the genomic sequence encoding *POU1F1* beta isoform in mammals and chicken. **E**. Exon trapping assay with pSPL3 exon trap vector containing exon 2 of *POU1F1* and portions of the flanking introns. **F**. Ethidium bromide-stained gel of exon trap products from cells transfected with the indicated plasmid. Arrowheads indicate the expected products for exon skipping (Blue), alpha isoform (Yellow), and beta isoform (green). **G**. *POU1F1* minigenes spanning from intron 1 to 5, with all of the intervening exons, were engineered with the indicated variants and assayed for splicing. WT and p.76L *POU1F1* splice to produce the alpha isoform, the G>T change in the splice acceptor causes exon skipping (red arrow) and the other patient variants all splice to produce *POU1F1* beta isoform.

### Patient missense variants disrupt normal *POU1F1* splicing to favor the beta isoform

Alpha is normally the predominant *POU1F1* isoform, but its splice acceptor is predicted to be much weaker than the beta isoform acceptor 78 bp upstream (MaxEntScan^42^; scores, alpha: - 3.63, beta: 6.96) (**Fig. 2C**). The beta isoform splice acceptor sequence and coding region are evolutionarily conserved in mammals and birds (**Fig. 2D**). We reasoned that splice repressor and/or enhancer sequences in *POU1F1* may dictate the normal balance of alpha over beta isoforms, and these may be disrupted by the four patient T>G transversions. To test the effect of these variants directly, we cloned *POU1F1* exon 2 and portions of the flanking introns into the exon trap splice reporter pSPL3 and introduced each variant by site directed mutagenesis (**Fig. 2E**). These small minigenes were transfected into COS-7 cells, and RNA was analyzed by RT-PCR. The wild type minigene produced almost exclusively alpha isoform, while each of the patient variants predominantly produced the beta isoform (**Fig. 2F**). We also tested splicing with larger minigenes, which contain portions of intron 1 and intron 5 with intact exons 2, 3, 4 and 5 as well as introns 2, 3 and 4, and obtained similar results, indicating the additional sequence context does not strongly influence the observed splicing pattern (**Fig. 2G**). We also tested two previously reported *POU1F1* variants in the longer minigene context. The c.214+1 G>T caused skipping of exon 2, as expected, resulting in an in-frame POU1F1 protein that lacks 80% of the transactivation domain^43^. This variant is associated with mild hypopituitarism. The p.P76L variant is located in the transactivation domain, enhances POU1F1 interaction with other proteins, and is associated with severe, dominant IGHD ^24^. The effect of this variant on splicing had not been assessed previously, and we found that it produced predominantly alpha isoform expression, indistinguishable from wild type.

### Saturation mutagenesis screen for splice disruptive effects

We set out to systematically identify splice disruptive variants in *POU1F1* exon 2 using a massively parallel splice reporter assay. We designed oligonucleotide pools containing every possible single nucleotide variant across exon 2 (150 bp) and 210 bp of the flanking introns (N*=*1080), and generated libraries in which these allelic series replaced the wild type *POU1F1* fragment in the pSPL3 reporter. To track the splicing outcomes associated with each mutation, we placed a degenerate 20mer barcode in the downstream 3’ UTR. The mutant plasmid library was subjected to subassembly sequencing^29^ to establish the pairing between each unique barcode and its associated *POU1F1* mutation. In total, the mutant library contained 255,023 distinct barcoded clones, among which 188,772 (74.0%) had exactly one programmed mutation. Nearly every targeted mutation appeared in this library (1070/1080, 99.1%), with a high degree of redundancy (median 75.0 distinct barcodes/mutation, **Suppl. Fig. 2**).

The splice reporter library was transfected as a pool into COS-7 cells and processed similarly to the single mutation constructs. Spliced reporter transcripts were read out *en masse* using paired-end RNA-seq (**Fig. 3A**), with each forward read measuring an individual splicing outcome and the paired reverse read containing the 3’ UTR barcode which identifies the mutation(s) present in the primary transcript. We performed 14 biological replicates, across which 94.2% (81.8-93.4%, mean 87.4%) of clone library single nucleotide variant associated barcodes were detected. As expected, alpha was the predominant *POU1F1* isoform (69.2% of reads overall), followed by exon 2 skipping (25.6%), and beta (1.6%). We created a catch-all category (‘Other’) for the remaining reads (3.6%) derived from the 262 other isoforms detected. Most of those noncanonical isoforms were only scarcely used; among them, the top 20 accounted for >80% of the reads from that category. For each *POU1F1* variant, a percent spliced in (PSI) value was computed for each isoform (alpha, skip, beta, other), averaged over the associated barcodes. PSI values were highly reproducible across replicates (median pairwise Pearson’s *r:* 0.92; **Suppl. Fig. 3**).

**Figure 3.**
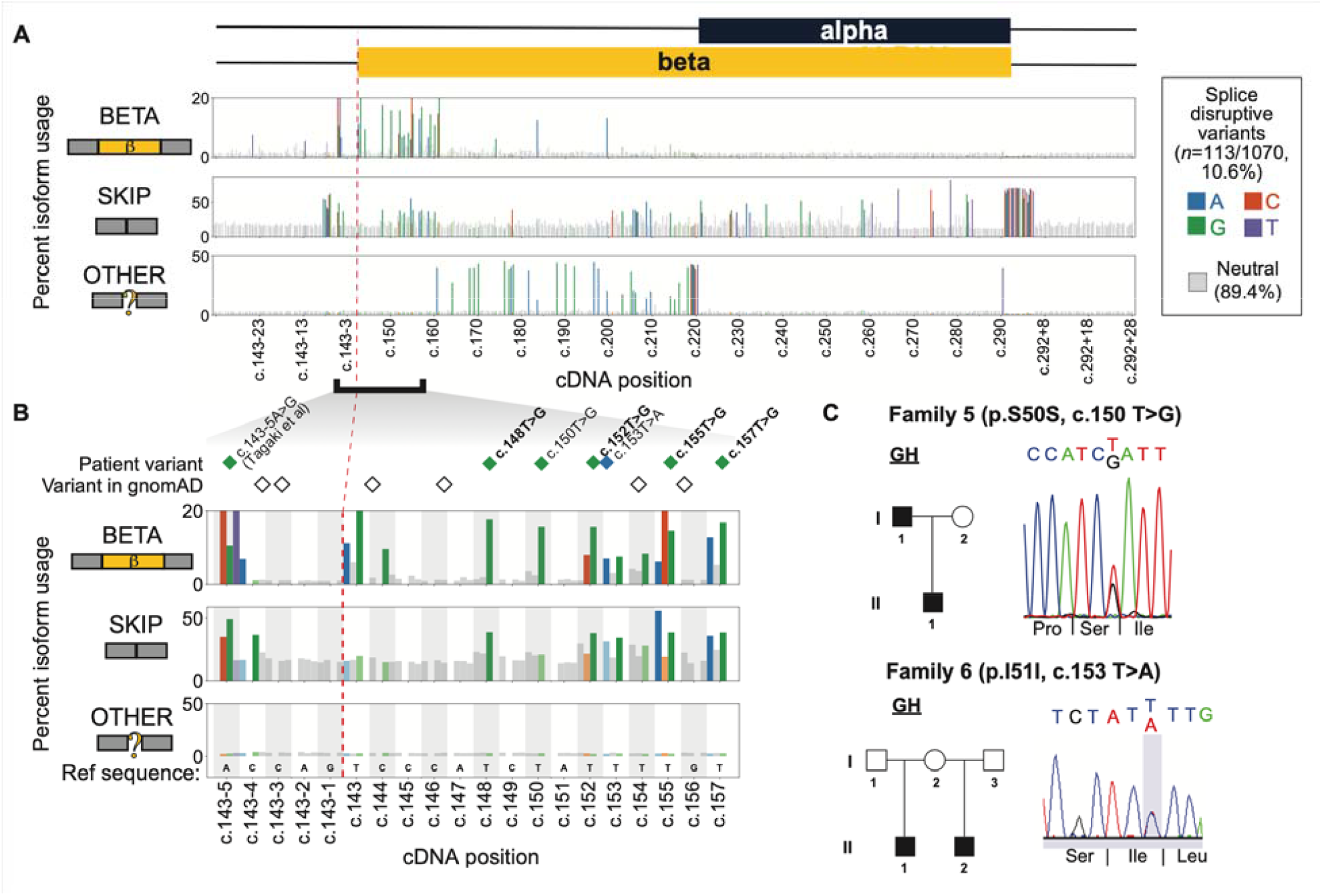
Splicing effect map in *POU1F1* exon 2 and flanking introns, and identification of IGHD families with synonymous changes. **A**. Percent usage of *POU1F1* exon 2 beta (top panel), skip (middle), and other isoforms (bottom) by variant position as measured by massively parallel minigene assay. Gray bars denote splicing-neutral variants, while colored bars indicate the base pair change of each SDV. Cropped intronic regions are shown in **Suppl. Fig. 4**. **B**. A cluster of SDVs near the beta isoform splice acceptor leads to increased usage of the beta isoform, and in some cases, increased exon skipping. Diamonds colored by the alternate allele indicate patient variants, and empty diamonds indicate variants reported in gnomAD. Missense variants’ labels are in bold text. **C**. Families 5 and 6 each had two individuals affected with IGHD and synonymous variants that were splice disruptive. Pedigrees and Sanger sequence confirmation of variants are shown.

### Splice disruptive variants (SDVs) across *POU1F1* exon 2

We assessed the splicing effect of 1,070 single nucleotide variants (**Figure 3A** and **Supp. Fig. 4**). Of these, 113 (10.6%) were splice disruptive variants (SDVs), which we defined as those which increased usage of beta, skip, or other isoforms by _≥_1.5 standard deviations. SDVs leading to increased skipping were the most frequent (*n* = 69/113; 61.1%), followed by those creating other isoforms (*n* = 34/113; 30.1%), and those which increased beta usage (*n* = 29/113; 25.7%), with some variants (*n* = 19/113; 16.8%) impacting usage of multiple isoforms (**Suppl. Fig. 5**). Variants leading to each outcome tended to cluster in distinct regions; notably, the beta-increasing SDVs were located near the 5’ end of the beta isoform. Intronic SDVs tended to lead to skipping. A few variants that increased skipping were scattered across exon 2, and there was some enrichment in the 5’ end of the beta isoform coding region, but most were enriched near splice donor and acceptor sites: 27 of 28 intronic SDVs were within +/- 20 bp of exon 2.

SDVs which created novel (“other”) isoforms were nearly all located within the coding region unique to the beta isoform and at the alpha isoform acceptor site (*n* = 33/34; 97.1%). Of these, most (27/34) create a novel acceptor AG dinucleotide that outcompetes the more distal, native alpha acceptor (**Suppl. Fig. 6**). Most of these (19/27) result in a frame-shifted transcript, and all are predicted to undergo non-sense mediated decay due to premature termination codons >50 bp upstream of the final exon junction. In contrast, each of the six mutations in the alpha isoform acceptor dinucleotide activates another cryptic acceptor six bp downstream, leading to in-frame deletion of two codons (**Suppl. Fig. 7**). Out of 99 mutations creating a GT dinucleotide, only one was used as a novel splice donor, c.290:C>T located 4 bp upstream of the native donor.

We compared these SDVs to the variants found in Families 1-4. All four patient missense variants showed strongly increased beta isoform usage (beta isoform z-scores range: 5.12-6.18), mirroring the results of individual minigene assays (**Fig. 3B**). Our results also recapitulate previously described effects of two variants found in CPHD patients: first, an upstream intronic variant c.143-5:A>G^44^ which led to increased beta usage and skipping (beta isoform *z*=3.48, skip *z* =3.13; **Figure 3C**), and an essential splice donor variant c.292+1 G>T which led to near-complete skipping (skip *z*=5.05; **Suppl. Fig. 7**)^43^.

We also examined the incidence of splice disruptive *POU1F1* variants in the general population. The gnomAD database contains 93 of the variants measured here; among those, eight (8.6%) are splice disruptive, all which are individually rare (minor allele frequency _≤_1.6×10^−5^; **Suppl. Fig. 8**). Overall, variants found in gnomAD were not significantly depleted for splice disruptive effects relative to randomly selected subsets of the tested single nucleotide variants (*p=*0.60, Fisher’s Exact Test). Thus, *POU1F1* SDVs are tolerated to a similar extent as other predicted loss of function variants (stop gain, frameshift, splice site), which are observed throughout *POU1F1* at low frequencies in gnomAD.

### Identification of a silent variant in *POU1F1* in a hypopituitarism patient

We next examined the splicing impacts of synonymous variants, which would typically be given low priority during genetic screening due to their expected lack of coding impact. Of the 111 synonymous variants tested, 23 were splice disruptive (20.7%; **Suppl. Fig. 5**). We identified unrelated patients with IGHD carrying two of these synonymous SDV in the beta isoform coding region near the 5’ end of exon 2 (**Fig. 3C**), both of which were absent in gnomAD and population-matched control databases. The first, c.150T>G (p.Ser50=), was found among an Argentinian cohort (*n*=171) in a family with two individuals with severe short stature and IGHD (**Table 1**), for whom WES did not reveal any likely pathogenic variants in known CPHD or IGHD genes. The index case had pituitary hypoplasia, and the patient responded well to recombinant GH treatment. The second, c.153T>A (p.Ile51=), was found in a French family in relatives with severe IGHD. The parent’s DNA was not available for testing, and the parent could be an unaffected carrier or an example of gonadal mosaicism. Each of these two silent variants increased beta isoform usage (beta *z*=5.33 for c.150T>G; beta *z*=1.79 for c.153T>A) to a degree similar to that of the four patient missense variants.

### Comparison to bioinformatic splicing effect predictions

We examined how scores from splicing effect prediction algorithms compared with these experimental measurements. We scored each single nucleotide variant in the targeted region of *POU1F1* using SpliceAI^35^, MMSplice^36^, SPANR^34^, HAL^33^ and ESRseq scores^37^. Among these, only SpliceAI predicted a high density of SDVs specific to the exon 2 beta region surrounding the patient variants (**Suppl. Fig. 9**). To benchmark each bioinformatic predictions, we took our SDV calls as a truth set and computed for each algorithm the area under the precision recall curve (**Suppl. Fig. 10**). SpliceAI was the most highly concordant with our results for both exonic variants (prAUC=0.790 vs other tools’ range: 0.329-0.430) and intronic variants (prAUC=0.661 versus other tools’ range: 0.526-0.604). Nevertheless, SpliceAI disagreed with our measurements for numerous variants: at the minimum threshold needed to capture all six patient variants as disruptive (SpliceAI deltaMax score_≥_0.19), it achieved 67.3% sensitivity (n=37 SDVs not predicted by SpliceAI) and 97.2% specificity (n=27 variants predicted by SpliceAI but not identified by our assay) for predicting the SDVs we identified. The degree of concordance with SpliceAI was largely insensitive to the specific measurement threshold used to call variants as splice disruptive in our screen (**Suppl. Fig. 10**). Additional studies will be required to resolve the discordant predictions for variants observed during clinical screening.

## Discussion

We found six unrelated cases with CPHD or IGHD that can be explained by variants that shift splicing to favor the repressive beta isoform POU1F1. The missense variants, p.S50A, p.I51S, p.L52W, and p.S53A, retain repressive function. They act in a dominant negative manner by suppressing the ability of the POU1F1 alpha isoform, expressed from the wild-type allele, to transactivate expression of *POU1F1* and other downstream target genes. Using saturation mutagenesis coupled to a high-throughput RNA-seq splicing readout, we systematically tested nearly every possible single nucleotide variant in or near *POU1F1* exon 2 for splice disruptive potential (**Supplemental Table 1**). We identified 113 SDVs which similarly activate usage of the beta isoform or cause other aberrant splicing outcomes such as exon skipping.

This screen accurately identified the four missense variants we identified in patients. It also identified 23 synonymous splice disruptive variants, or over one-fifth of the possible synonymous variants in *POU1F1* exon 2. We identified two of these synonymous SDVs in unrelated families with IGHD, c.150T>G (p.Ser50=) and c.153T>A (p.Ile51=), each of which increased beta isoform usage similarly to the four patient missense variants that initially drew our attention. These findings underscore the need to closely examine variants for splice disruptive effects, particularly synonymous variants that could be overlooked by traditional exome sequencing filtering pipelines.

The clinical features varied amongst the six families, although they were consistent within a family. Families 1, 3, and 4 presented with CPHD, while Families 2, 5, and 6 had IGHD. Moreover, Family 4 developed hypocortisolism. The reason for this variability in presentation is unknown. However, there are precedents for variable clinical features and incomplete penetrance with other cases of hypopituitarism^18^. Approximately 50% of IGHD progresses to CPHD, and this can even occur when the mutated gene is only expressed in GH-producing cells, i.e. *GH1*^45^. Even individuals with the same *POU1F1* mutation (i.e. p.E230K) can present with either IGHD or CPHD^46^, indicating a contributing role for genetic background, epigenetic, and/or environmental factors. Both affected relatives in Family 1 had stalk disruption, a phenotype not currently associated with any other *POU1F1* variants. This feature may be due to the presence of an additional variant in *SIX3*, p.P74R, that was carried by two unaffected relatives. Heterozygous loss of function of *SIX3* is associated with incompletely penetrant and highly variable craniofacial abnormalities, including CPHD and holoprosencephaly, and there is precedent in mice for *Six3* loss of function to exacerbate the phenotype caused by mutations in other CPHD genes such as *Hesx1*^47-49^.

Autosomal dominant inheritance is clear in Family 2, in which there were four affected individuals over three generations, as well as Families 4, 5, and 6. POU1F1 acts as a heterodimer^50^. Some other dominant mutations in *POU1F1* act as negative effectors due to the ability of the mutant protein to interfere with the action of the wild type protein produced from the other allele^25,51,52^. The negative effect of POU1F1 beta on the transactivation properties of POU1F1 alpha are context dependent, with differential effects on *Gh, Prl* and *Pou1f1* reporter genes^53^. The strongest effect was reported for autoregulation of POU1F1 expression via the distal, late enhancer; dampening the auto-activation of *POU1F1* expression, and adversely affecting differentiation of the entire POU1F1 lineage and result in anterior lobe hypoplasia.

The lack of significant depletion for *POU1F1* SDVs among ostensibly healthy adult populations underscores the possibility of variable expressivity and/or penetrance for *POU1F1* splice-disruptive variants. This is consistent with the apparently unaffected parents in Families 1 and 6. A subset of these variants, like c.222T>C which disrupts the alpha isoform acceptor and causes a frame-preserving two-codon deletion, may retain partial or complete function. Still others, may cause loss-of-function without dominant negative effects, and would not be expected to be strongly depleted.

In human genes, canonical splice site motifs contain less than half of the information content needed for proper splicing ^54^. Additional specificity is provided by short (6-10 nt) motifs termed exonic or intronic silencers and enhancers, which are bound by RNA binding proteins that promote or antagonize splicing^55^. Although transcriptome-wide atlases have been developed to map these sites^37,56^, and derive motif models^57^, it often remains unclear how genetic variants impact their binding and in turn the eventual splicing output. Our splicing effect map identifies a cluster of SDVs at the 5’ end of the *POU1F1* exon 2, each of which increases the usage of the normally repressed beta isoform. These results suggest the presence of an exonic splice silencer (ESS) which may normally suppress utilization of the beta isoform acceptor. We mined the cisBP-RNA database ^38^ and identified nine candidate motifs with strong matches to the U-rich wild-type sequence in this region (c.143-1 to c.167) corresponding to known splicing factors including ELAVL1 (HuR), RALY, TIA1, and U2AF2 (**Suppl. Fig. 11**). All six patient variants replaced a U with another base (G in 5 of 6 bases), which may disrupt these motifs at high information content positions (**Suppl. Fig. 12**). Other variants predicted to disrupt these motifs tended to be beta-promoting more often than neutral in our map (*p* < 0.01, Fisher’s Exact test). These trends suggest that U-rich ESS serves to inhibit production of POU1F1 beta and this inhibition is disrupted by CPHD-associated variants, although conclusively identifying the specific cognate binding factor will require further study.

These results extend the breadth of endocrine disorders caused by disrupted splicing. For example, in a large cohort with IGHD from Itabaianinha, Brazil, affected individuals are homozygous for a mutation in the splice donor dinucleotide (c.57 + 1G > A) in the growth hormone releasing hormone receptor gene (*GHRHR*)^58^. In addition, most mutations that cause dominant IGHD type II affect splicing of the growth hormone (*GH1*) gene^59^. Mutations in splice sites or splice enhancer sequences result in skipping exon 3 and production of a dominant-negative 17.5 kD isoform of growth hormone that lacks amino acids 32-71^60^. The severity of the disease is variable and correlates inversely with the ratio of 17.5 to 20 kD GH. Finally, severe short stature associated with Laron syndrome, or GH resistance, can be caused by generation of a cryptic splice site in the GH receptor gene. Individuals from El Oro and Loja in southern Ecuador are homozygous for a p.180E codon variant (GAA to GAG) that do not change the amino acid encoded but create a splice acceptor site 24 nt upstream of the normally utilized site^61^. It is notable that antisense oligonucleotide therapies hold promise for treating diseases caused by abnormal splicing, including IGHD^62,63^.

Splicing disruption accounts for a significant minority of the genetic burden in endocrine disorders, as in human genetic disease more generally ^64,65^. Some estimates from large-scale screens indicate that 10% of SNV within exons alter splicing, and a third of all disease associated SNVs impact splicing efficiency^66^. Variants at or near canonical spice sites are readily recognized as pathogenic^67^, and these can be identified predicted with high accuracy by algorithms such as SpliceAI. However, for exonic variants, particularly those farther from exon junctions, splicing defects may be more challenging to identify bioinformatically^68-70^. Efforts to interpret these variants will need to account for the functional impacts of changing the encoded protein sequence as well as its splicing. Finally, as our results illustrate, different variants in a single gene may lead to distinct splicing outcomes with diverse consequences ranging from the straightforward loss-of-function to dominant negative effects.

## Data Availability

Data will be posted on GEO and Github

## Acknowledgements

This work was supported by the National Institutes of Health (R01HD097096 to SAC), the Japan Society for Promotion of Science (HB), Grant 2013/03236-5 from the São Paulo Research Foundation (FAPESP) (IJPA), a grant from Pfizer (RP), and the Argentinean National Agency of Scientific and Technical Promotion, PICT 2016-2913 and PICT 2017-0002 (MIPM).

## Author Contributions

<u>Transfection and exon trapping</u>: Peter Gergics

<u>Families 1, 3 and 6 patient collection and candidate gene screening</u>: Frederic Castinetti, Frédérique Albarel, Alexandru Saveanu, Anne Barlier, Thierry Brue

<u>Family 2 patient collection and candidate gene screening</u>: Alexander Jorge, Luciani Renata Silveira Carvalho, Marilena Nakaguma, Berenice B Mendonça, Ivo JP Arnhold

<u>Family 4 patient collection and WES</u>: Denise Rockstroh-Lippold, Julia Hoppmann, Rami Abou Jamra, Roland Pfaeffle

<u>Family 5 patient collection:</u> Debora Braslavsky, Ana Keselman, Ignacio Bergadá <u>Family 1 WES and analysis:</u> Qing Fang, A Bilge Ozel, Qianyi Ma, Jun Z. Li <u>Family 2 WES and analysis:</u> Michael H. Guo, Andrew Dauber

<u>Family 5 WES and analysis</u>: Sebastian Vishnopolska, Julian Martinez Mayer, Marcelo Martí, María Ines Pérez Millán

<u>High throughput mutagenesis and analysis</u>: Cathy Smith, Mariam Maksutova, Jacob O. Kitzman <u>Wrote manuscript</u>: Hironori Bando, Cathy Smith, Jacob O. Kitzman, Sally A. Camper

## Supplementary Figures and Legends

**Supplementary Figure 1.**
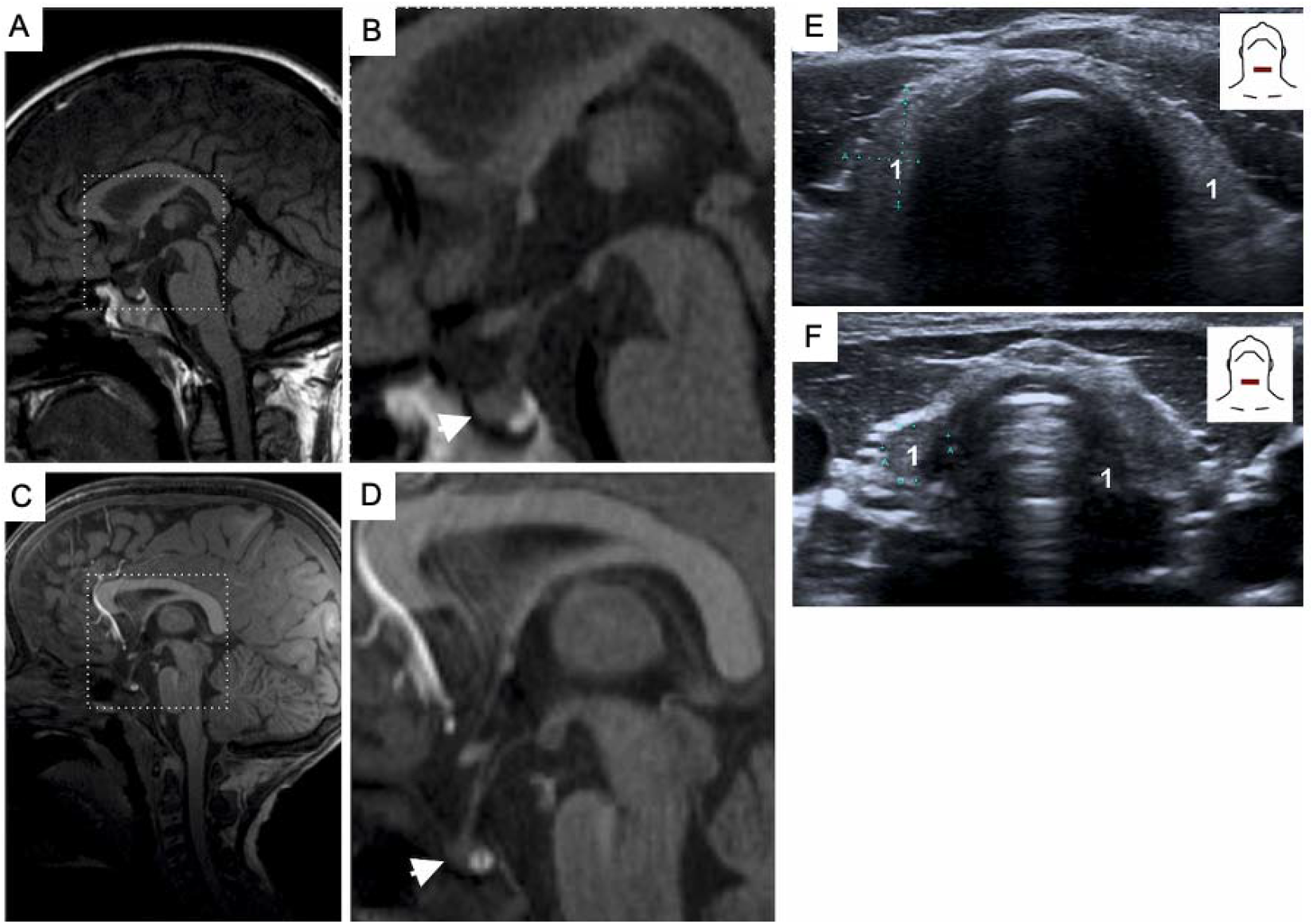
Clinical information for Family 4. Brain MRI of patient I.1 (**A, B**; as a teenager) and II.1 (**C-D**; as a pre-teen). Thyroid ultrasound of patient I.2 (**E**) and II.1 (**F**). ^1^Arteria carotis communis.

**Supplementary Figure 2.**
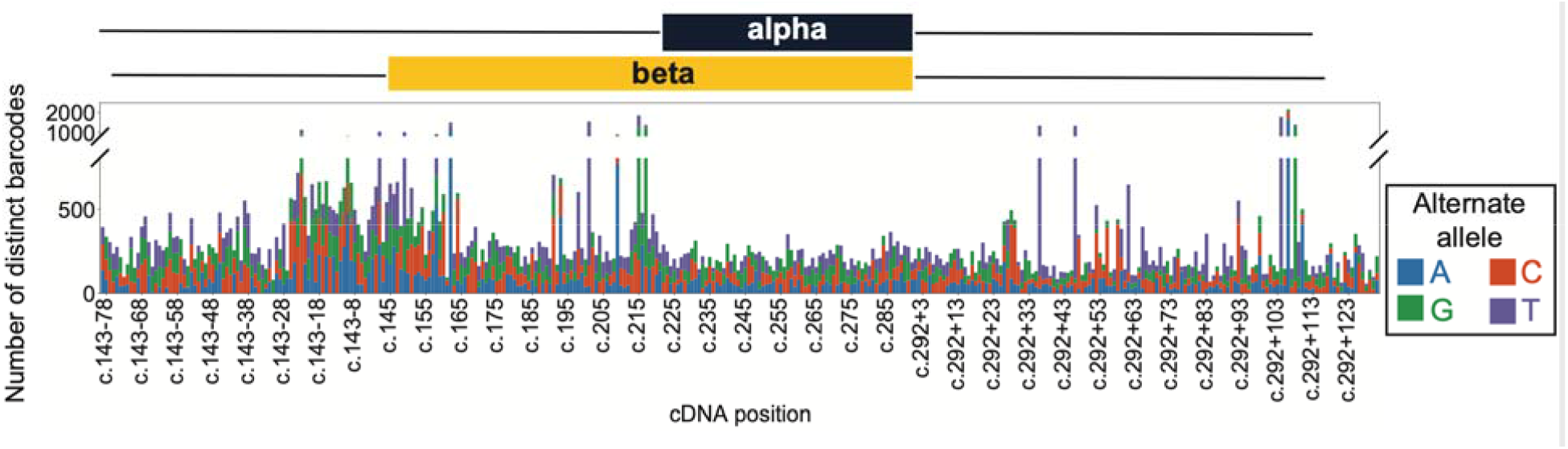
Completeness and uniformity of saturation mutagenesis. Stacked barplot showing, for each *POU1F1* variant by position (x-axis) and allele (color), the number of distinct barcodes detected in RNA-seq data (median across replicates).

**Supplementary Figure 3.**
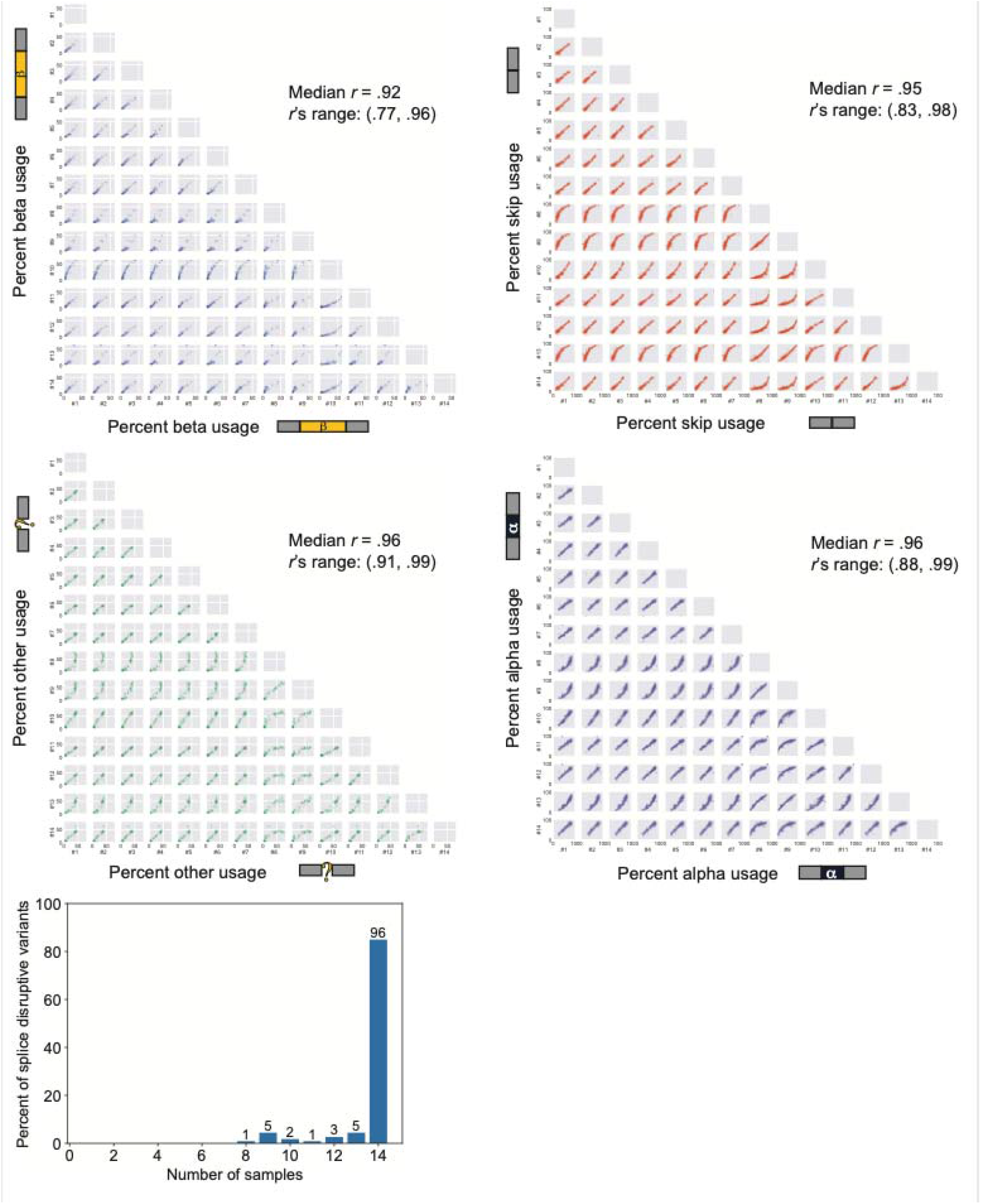
Inter-replicate correlation. A. Pairwise scatterplots of percent isoform use for beta, skip, other, and alpha isoforms among the fourteen biological replicates. The median and range of Pearson’s correlation values across samples are shown for each isoform. B. Histogram plotting the number of replicate samples in which variants met the splice disruptive variant (SDV) criteria; all SDVs met threshold in _≥_8 replicates, with 96/113 found in all 14 replicates.

**Supplementary Figure 4.**
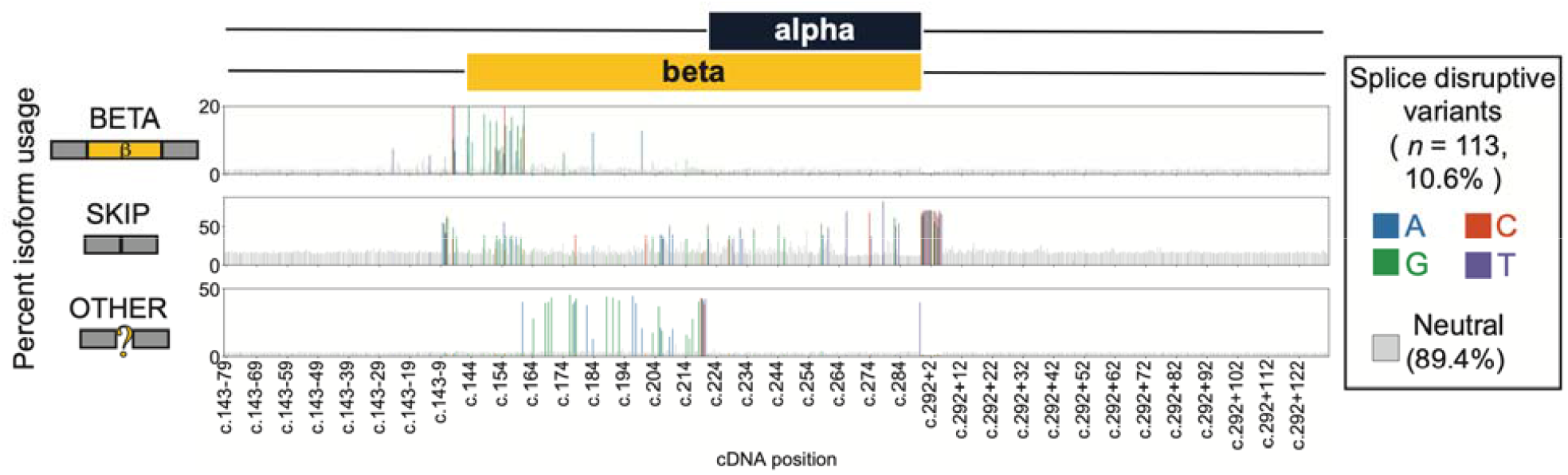
Uncropped *POU1F1* splicing effect map. Uncropped version of **Figure 3A**, including cropped intronic regions lacking any SDVs. Isoform usage and variants are plotted as in **Figure 3A**.

**Supplementary Figure 5.**
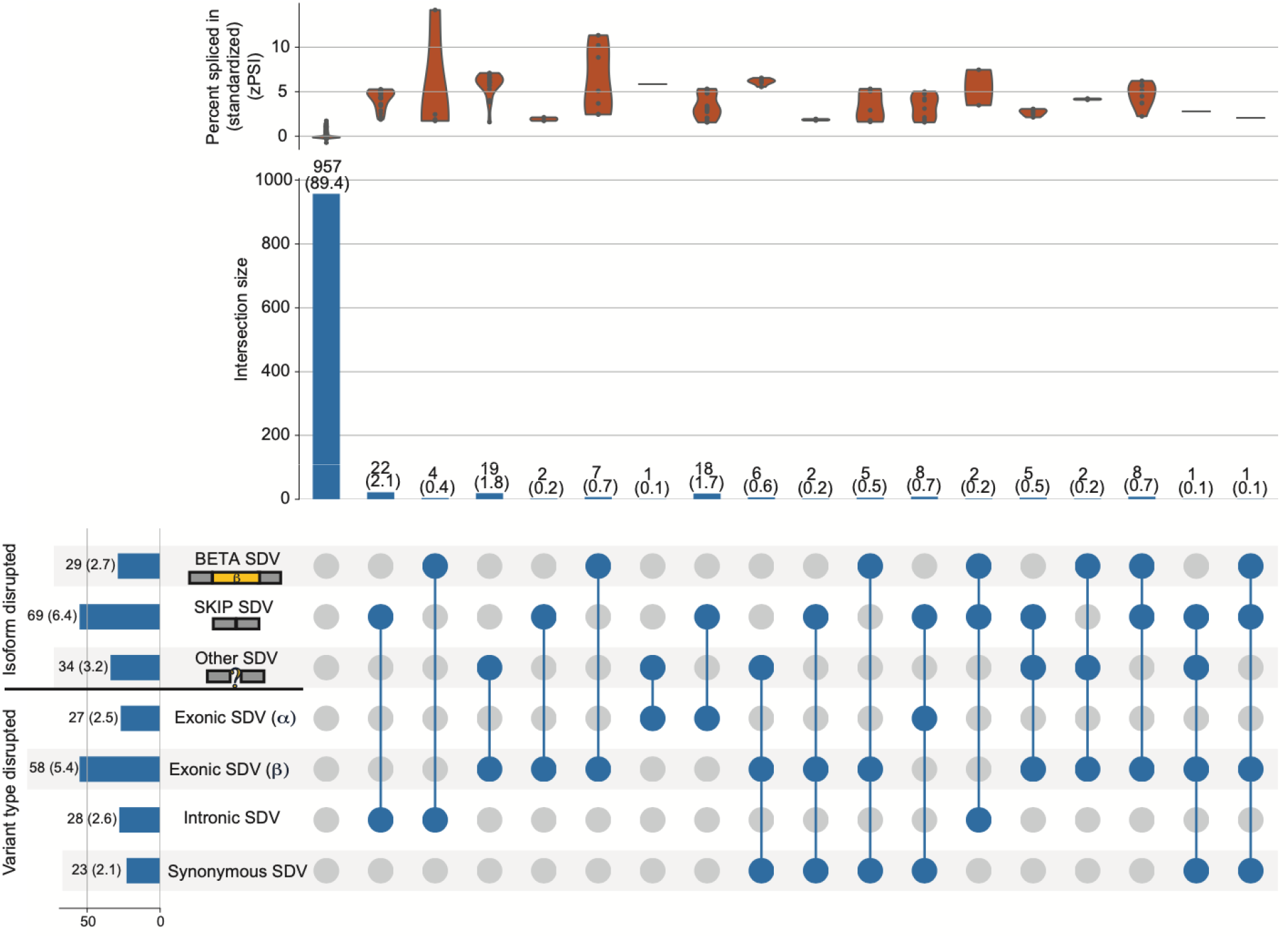
Splice disruptive variants by isoform and variant type. Distributions of isoform usage *z*-scores for each subset of subsets are shown as violin plots. Count within each intersection (and % of total) are shown above vertical bars. Count within each subset prior to intersection (and % of total) are shown along horizontal bars. UpSet plot showing splice disruptive variants (SDVs), categorized by isoform (beta, skip, and other) and variant type (exonic, intronic, and synonymous). Filled circles denote membership in multiple categories (e.g., second column from the left indicates there are 22 intronic SDVs causing increased skipping).

**Supplementary Figure 6.**
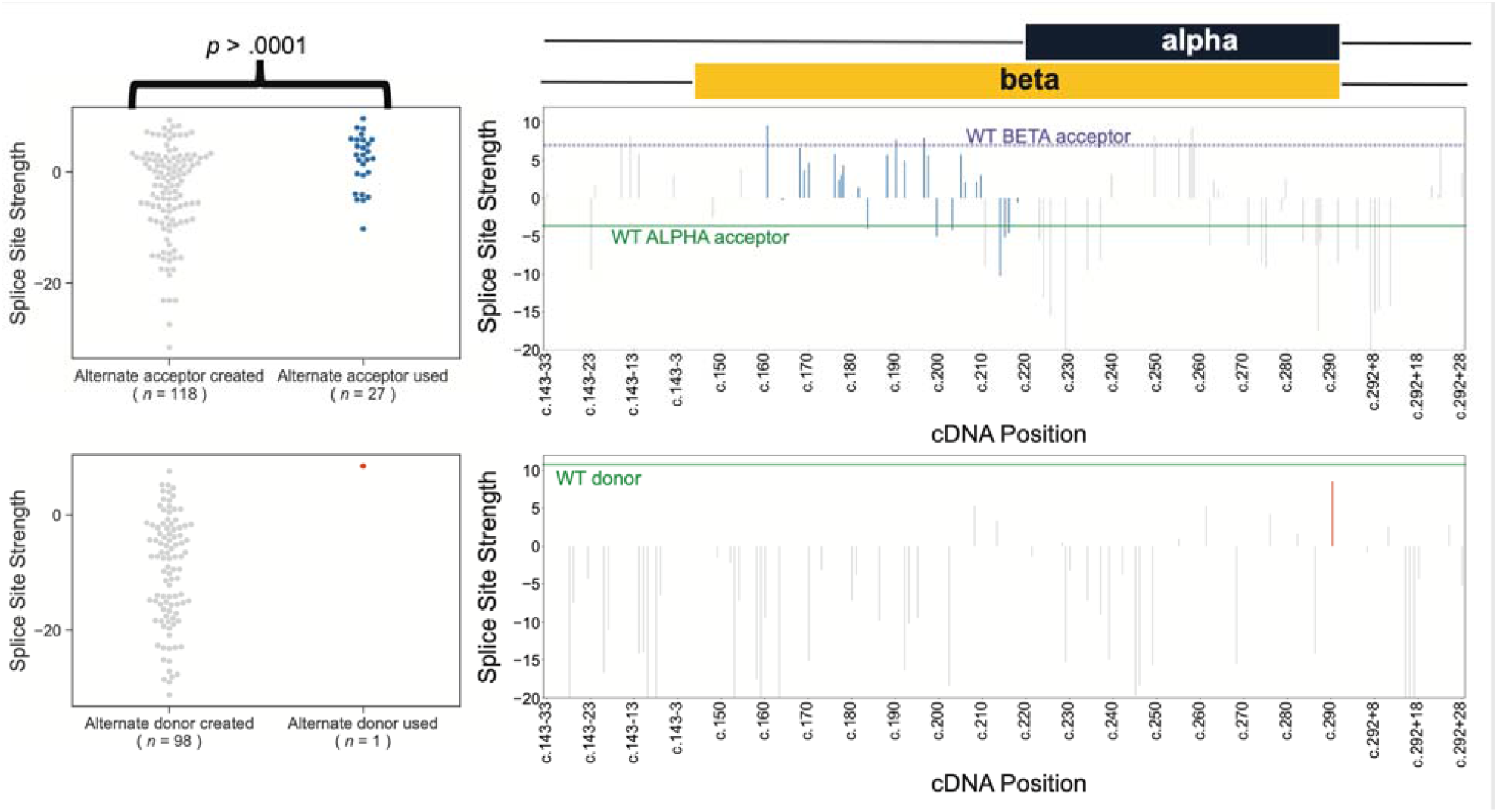
Splice site strength for novel alternate donors and acceptors. Splice site strength as predicted by MaxEntScan^42^ for novel alternate splice acceptor and donor sites. *P*-value corresponds to a *t*-test comparing the splice site strength for motifs at seldom used novel acceptor sites (*n* = 118) vs. motifs that promote use of another isoform at novel acceptor sites (*n* = 27). Dashed line (purple) represents the splice site strength of the native beta acceptor site. Solid lines (green) indicate the splice site strength of the native alpha acceptor site and native donor site respectively. Splice site strength is truncated at -20 in the positional plots, but minimum is as low as -31.6 for novel acceptors and donors within this exon.

**Supplementary Figure 7.**
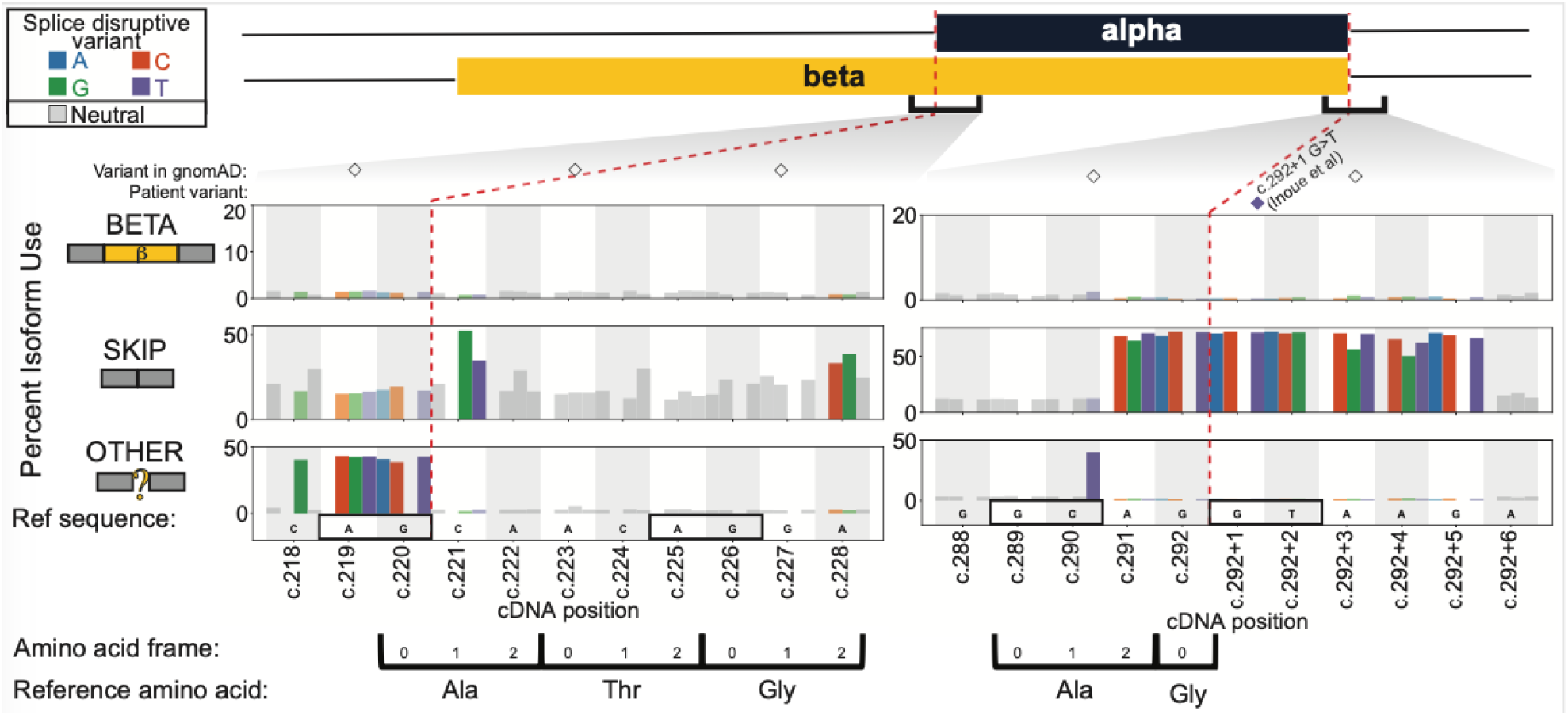
Alternate splice sites and frameshift mutations. Detailed view of splicing effect measurements, plotted as in **Figure 3B**, focusing on native alpha acceptor site (left) and native donor site (right). Colored and unfilled diamonds indicate patient variants (colored by alternate allele) and gnomAD variants, respectively. Canonical and cryptic splice sites are boxed, red dashed lines demarcate canonical exon boundaries, and coding frame and corresponding amino acids are indicated below.

**Supplementary Figure 8.**
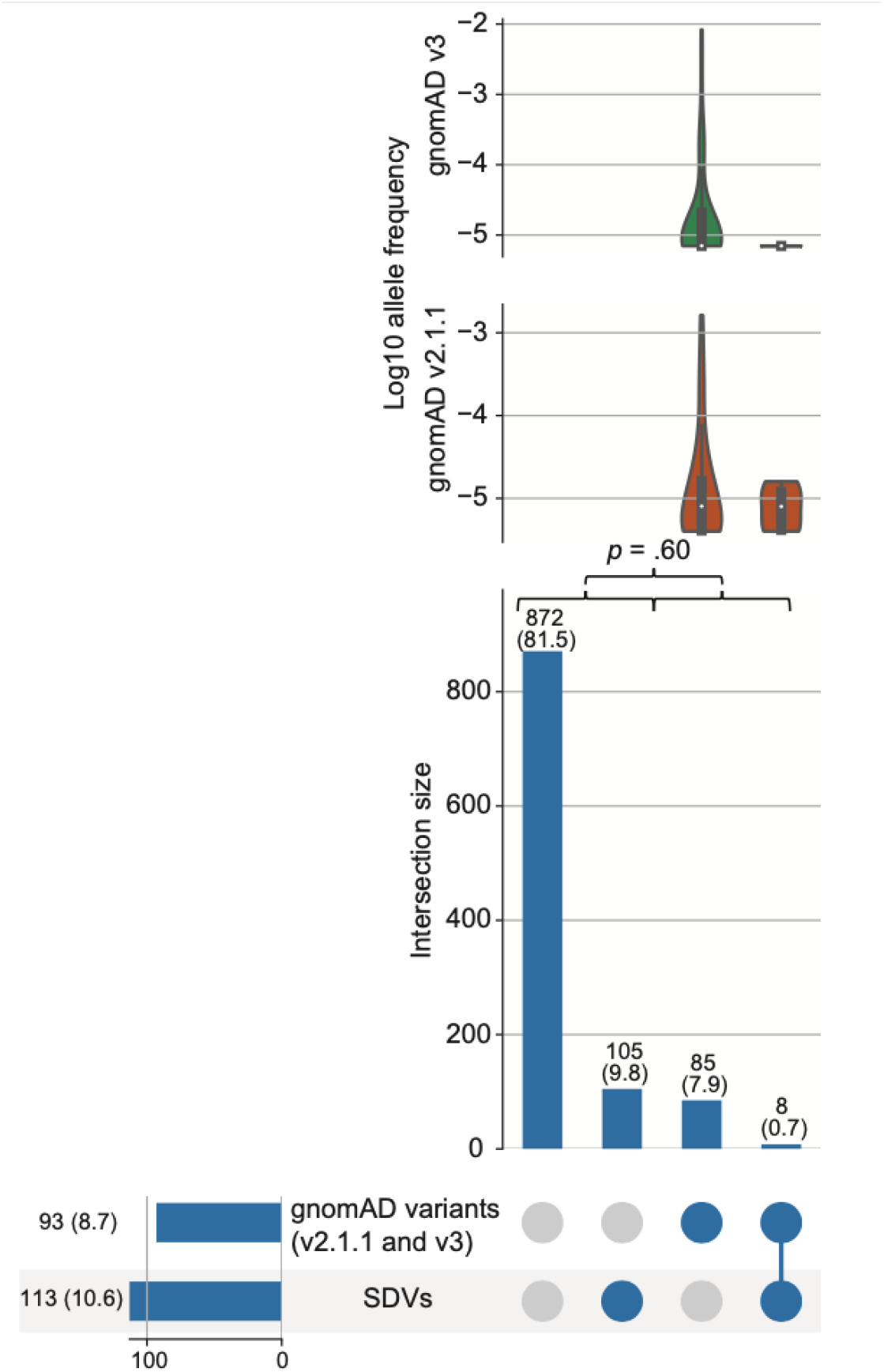
Splice disruptive variants (SDVs) in gnomAD. Violin plots of the log10 allele frequency for each variant found in gnomAD v2.1.1 (orange) and v3 (green) within each subset are shown. Count within each intersection (and % of total) are shown above vertical bars. Count within each subset prior to intersection (and % of total) are shown along horizontal bars. *P*-value corresponds to a Fisher’s exact test comparing the proportion of splice disruptive variants between gnomAD variants and variants absent from gnomAD. UpSet plots showing intersection of tested variants with or without splice disruptive effects, and gnomAD variants.

**Supplementary Figure 9.**
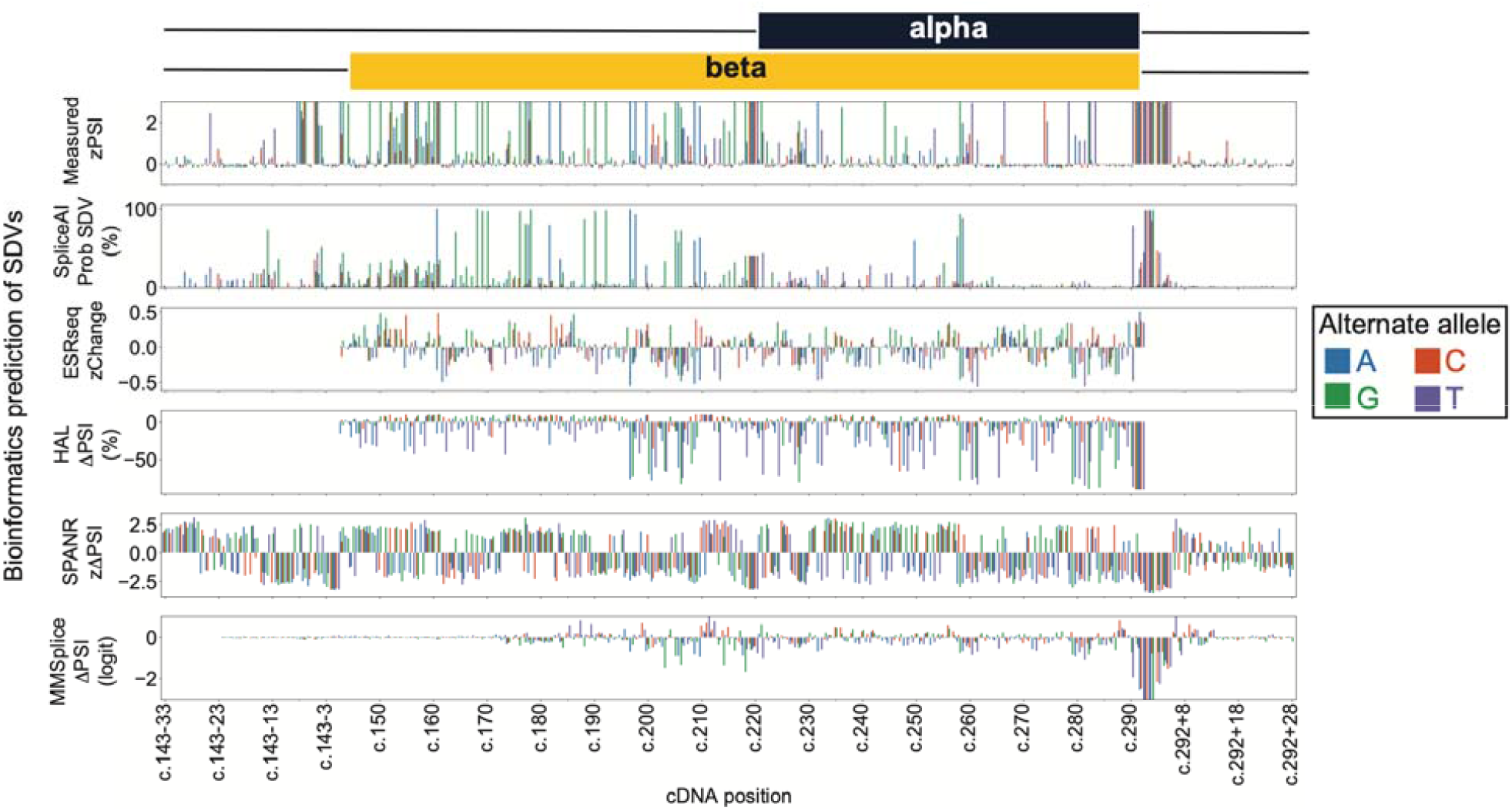
*In silico* predictions of splice disrupting variants (SDV). Barplots showing for each variant (color) at every position (x-axis) the splicing effect measurements (top y-axis) and splice disruption as predicted by SpliceAI, ESRSeq, HAL, SPANR, and MMSplice (from second from the top to bottom y-axes).

**Supplementary Figure 10.**
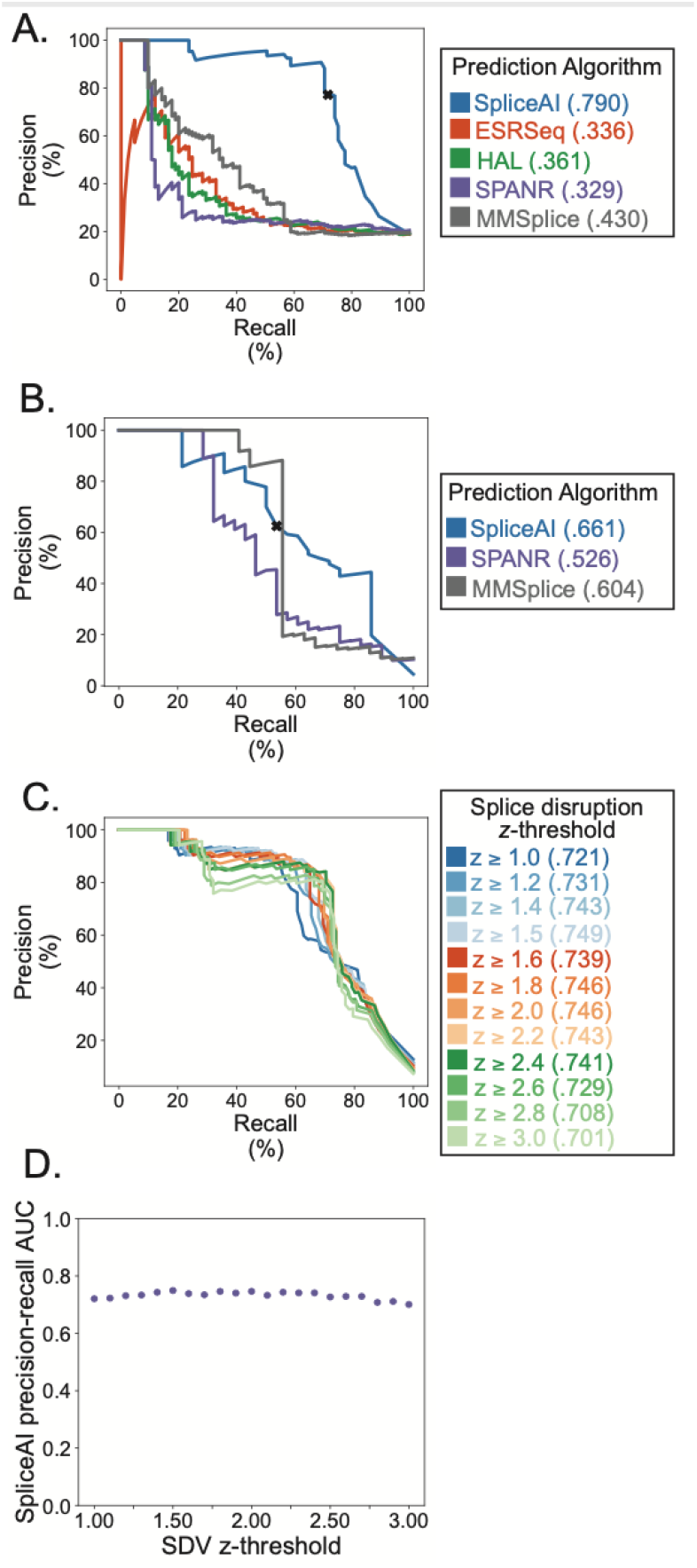
Evaluation of *in silico* splicing effect predictions. **A**. Precision-recall curve showing the precision (y-axis) and recall (x-axis) of SpliceAI (blue)^35^, ESRseq (orange)^37^, HAL (green)^33^, SPANR (purple)^34^, and MMSplice (gray)^36^ to predict splice disruptive variants (SDVs) in exonic regions. The ‘x’ is the at the minimum threshold where SpliceAI predicts all of the patient variants as disruptive (SpliceAI deltaMax score _≥_ 0.19). Area under the curve (prAUC) is shown within the legend **B**. Same as in A but for intronic variants. Since HAL and ESRseq values do not apply in noncoding regions so they are omitted from this plot. **C**. Precision-recall curve of the precision (y-axis) and recall (x-axis) for SpliceAI prediction of measured splice disruption across varying zPSI thresholds (range: 1 - 3) to call variants as disruptive. prAUC for each threshold is shown within the legend. **D**. Scatterplot of SpliceAI prAUC (y-axis) at varying splice disruption z-thresholds (x-axis).

**Supplementary Figure 11.**
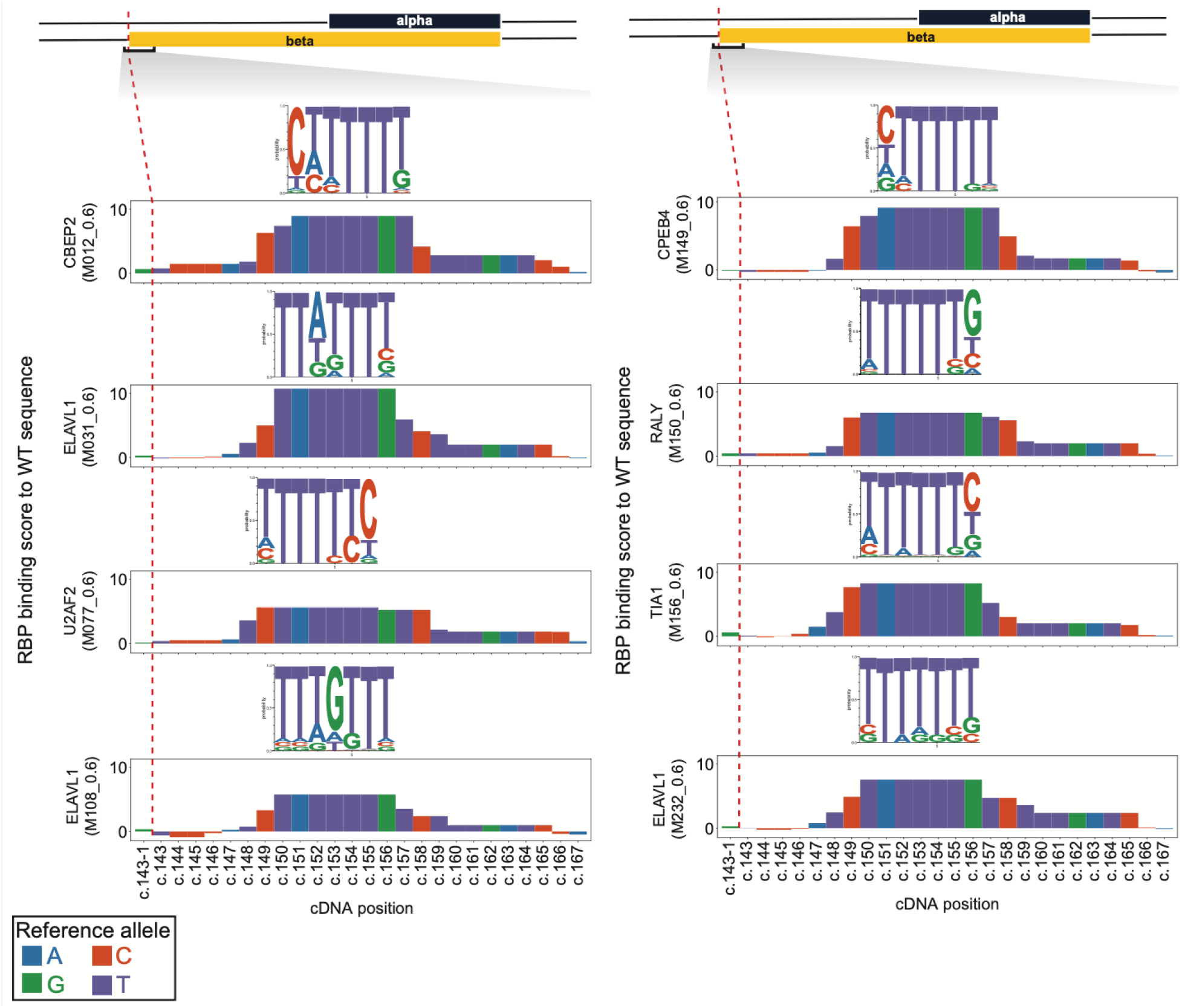
RNA binding protein consensus binding motifs relative to splice acceptor for *POU1F1* beta. Barplots displaying match scores (y-axis) for selected motifs defined by RNACompete^38^ scored against the wild-type *POU1F1* sequence beta region (positions c.143-1 to to c.167).

**Supplementary Figure 12.**
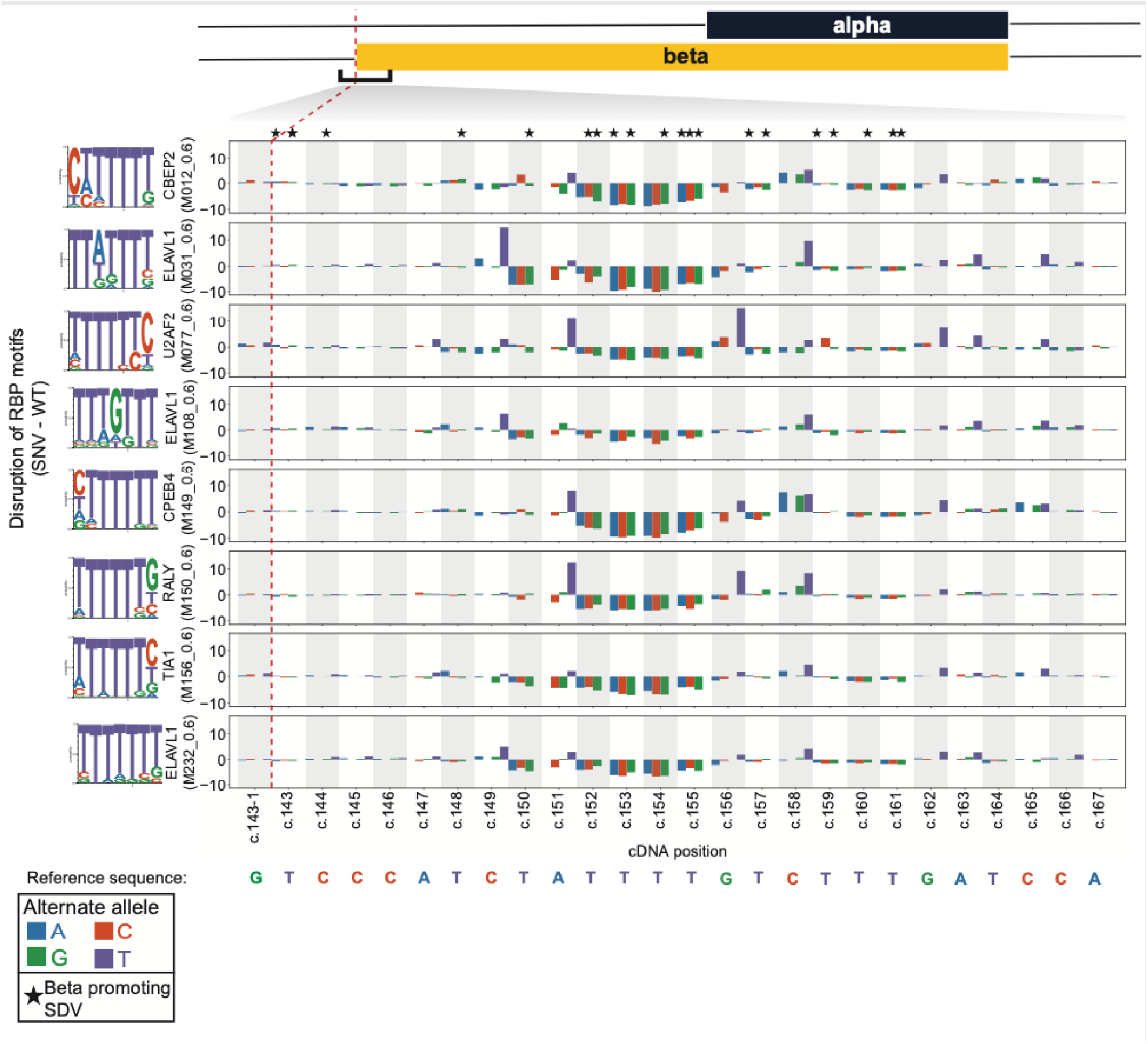
Changes in RNA binding protein motifs scores due to the SNVs in *POU1F1* beta. Barplots show the change in maximal RNAcompete kmer score (y-axis) by variant and position (x-axis), relative to the same motif scored against the wild-type *POU1F1* sequence. Black stars indicate SDVs that promote beta isoform splicing.

**Supplemental Table 1. Summary functional effects of all variants tested near *POU1F1* exon 2**.

